# Performance of a universally offered prenatal screening program incorporating cfDNA in Ontario, Canada: a descriptive population-based cohort study of 280,000 pregnancies

**DOI:** 10.1101/2020.09.22.20195123

**Authors:** Shelley D Dougan, Nan Okun, Kara Bellai-Dussault, Lynn Meng, Heather E Howley, Tianhua Huang, Jessica Reszel, Andrea Lanes, Mark C Walker, Christine M Armour

## Abstract

**Objectives:** To measure the population-based performance and impact of Ontario, Canada’s modified-contingent prenatal screening system for the detection of trisomies 21 (T21) and 18 (T18).

**Design:** A retrospective, descriptive cohort study examining routinely collected data from BORN Ontario, which captures linkable population data for prenatal and neonatal health encounters across a variety of settings (e.g., laboratories, birthing hospitals and midwifery practice groups).

**Setting:** A province-wide and publicly funded prenatal screening program in Ontario, Canada offering cfDNA screening for those at increased risk of having a pregnancy with T21 or T18.

**Participants:** 373,682 singleton pregnancies with an estimated due date between September 1 2016 and March 31, 2019 who were offered publicly funded prenatal screening.

**Main outcome measures:** Prenatal detection of T21 or T18, ascertained by cytogenetic results. Performance was assessed by calculating sensitivity, specificity, positive predictive value and negative predictive value against confirmatory diagnostic cytogenetic results and birth outcomes. The secondary objective was to determine the impact of contingent cfDNA screening by measuring uptake and the proportion of T21 screen-positive pregnancies undergoing subsequent cfDNA screening and invasive prenatal diagnostic testing (PND).

**Results:** 69% of pregnancies in Ontario underwent prenatal screening for T21/T18. The modified-contingent screen sensitivity was 89.9% for T21 and 80.5% for T18. The modified-contingent screen-positive rate was 1.6% for T21 and 0.2% for T18. The cfDNA screening test failure rate was 2.2% (final result including multiple attempts). The PND rate among pregnancies screened was 2.4%.

**Conclusions:** This study is the largest evaluation of population-based performance of a publicly funded cfDNA prenatal screening system. We demonstrated a robust screening system with high sensitivity and low PND consistent with smaller validation studies

## Introduction

The offer of screening for chromosomal aneuploidies, such as trisomies 21 (T21) and 18 (T18), is part of routine prenatal care internationally. Cell-free DNA (cfDNA) screening, introduced in Canada in 2013, significantly disrupted many standard screening programs [1–4]. Analyzing cell-free DNA originating from the placenta, this screening test is non-invasive, with better detection and fewer false positives than multiple marker screening (MMS)[5]. While the low false positive rate has been shown to translate into a reduced need for follow-up diagnostic testing, the price of the test (∼390 CAD)[6] has precluded it from being adopted into many publicly funded screening programs as a first-tier screening test. Given the option to self-pay for cfDNA screening, studies have shown that access is based on financial ability rather than medical need [7,8]. Internationally, there have been a variety of strategies for the equitable integration of cfDNA screening into prenatal screening protocols [5]. A common approach to optimize prenatal detection while maintaining cost-effectiveness is a contingent model, where cfDNA screening is offered for pregnancies at increased risk of aneuploidy [8], although the definition of ‘increased risk’ varies widely.

Evidence-informed policy on the adoption of new technologies and their impact on publicly funded programs requires system-wide data that is rigorously collected and robustly analyzed. Such evidence is lacking for prenatal screening, as analyses from many screening programs have been complicated by a lack of complete population ascertainment. Some are missing birth outcomes for *all* screened pregnancies [9][10], such that that specificity analyses cannot be performed. Other studies used fully modelled data [11], were enhanced with modeled data by extrapolating cohort findings into projected population performance [12], or used a screening and diagnostic testing cohort with unlinked estimated total births [13]. Performance of cfDNA screening has been published for numerous populations, but almost all are industry funded in part or in whole [14–17]. There is a paucity of data on complete population performance of either MMS or cfDNA screening within a system that captures sequential prenatal encounter data (including cytogenetic diagnosis and birth outcomes). To date, such data has only been made available from the ‘Danish Fetal Medicine Database’ [18,19] and ‘Victorian Perinatal Record Linkage (PeRL)’ study [13]. The Danish database has generated population-based performance data of first-trimester MMS from 2008-2012 for 268,342 singleton pregnancies [20], as well as contingent cfDNA screening (offered for pregnancies with a moderate risk result for trisomy 21 after first trimester MMS) from 6,449 singleton pregnancies[21]. The Victorian PeRL study used individual record linkage for 61,911 screened pregnancies to determine the utilization and population-based performance of different prenatal screening modalities. [13] Neither examined performance of a large population-based contingent screening model.

Ontario, Canada has provided publicly funded MMS for aneuploidies since the early 1990s, and has been a Canadian leader in the introduction of new prenatal screening technologies. In 2016, the Ontario government funded cfDNA screening for those that met specific high risk criteria, with the testing being performed in-province by two companies employing different technologies. In addition to a screen-positive MMS result, Ontario’s model includes several other criteria for accessing publicly funded cfDNA screening, as illustrated in Figure 1. Each of these companies contributes complete testing data to the Better Outcomes Registry &Network (BORN) Ontario (www.bornontario.ca), the province’s perinatal registry that collects information on pregnancy and newborn encounters with the health care system, including cytogenetic test results and birth outcomes. Within BORN Ontario, Prenatal Screening Ontario (PSO; https://prenatalscreeningontario.ca) is the government-funded program that coordinates provincial prenatal screening and facilitates the incorporation of evolving technologies and screening options (Supplemental Figure S1a). PSO is also responsible for ongoing quality assurance reporting; these activities are supported by the population data collected by BORN Ontario, using a variety of data assets from BORN Ontario (Supplemental Figure S1b).

**Figure 1:**
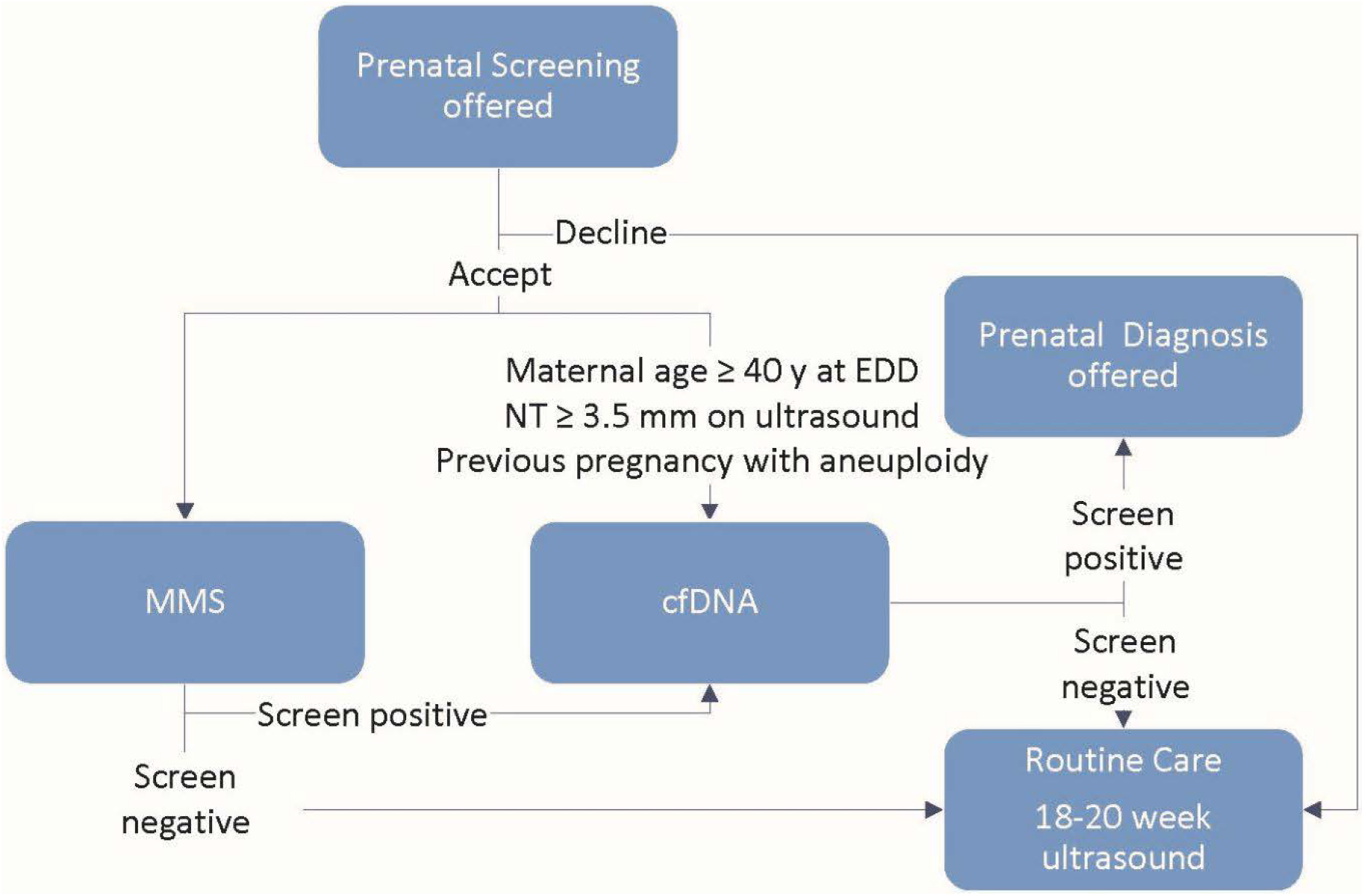
A Universal and Publicly Funded Modified-Contingent Model for Aneuploidy Screening in Ontario, Canada. In 2016, Ontario funded modified-contingent cfDNA prenatal screening for the common autosomal aneuploidies (trisomy 21, 18, 13) and sex chromosome aneuploidies. Women at high-risk for fetal aneuploidy are eligible for publicly funded cfDNA screening following screen-positive MMS (defined as risk ≥ 1 in 350 for enhanced first trimester screening or risk ≥ 1 in 200 for quad screening) or as a first-tier screen. Criteria for determining eligibility for first-tier cfDNA screening are based on recommendations from the provincial advisory group with the goal of optimizing performance while containing costs [30]. cfDNA: cell-free DNA screening; EDD: estimated due date; NT: nuchal translucency; MMS: multiple marker screening.

The primary objective of this study was to measure population-based performance (overall and modality-specific) of Ontario’s modified-contingent model of prenatal screening, incorporating publicly funded cfDNA screening for those at increased risk of having a pregnancy with T21 or T18. The secondary objective was to measure the impact of a contingent prenatal screening approach on subsequent screening and testing choices, including invasive prenatal diagnosis (PND).

## Methods

### Study Design

This was a retrospective population-based descriptive cohort study that examined routinely-collected data within BORN Ontario. Established in 2009, BORN Ontario is a prescribed registry under the Personal Health Information Protection Act (PHIPA) [22]. BORN Ontario collects critical health data about every pregnancy, birth and newborn in Ontario. Data are collected directly through a web-based platform from fertility clinics, prenatal screening and diagnostic laboratories, hospitals, midwifery practice groups and multiple other organizations across the province [23]. BORN Ontario uses ongoing data validation processes, quality assurance checks, and formal training sessions for individuals entering data to ensure high quality data capture [24,25]. Linkage of data elements and encounters across data sources is facilitated by a robust proprietary linking and matching algorithm, using unique provincial health numbers as the predominant identifiers and supplementing with other information to strengthen the linkage [25,26].

Direct upload and automated linkage of cytogenetic test records began in 2018. Cytogenetic records obtained before this date were matched using a multi-step deterministic matching strategy. All matched records were validated through visual inspection and an approximate 10% sample per cytogenetic laboratory were visually inspected.

The BORN registry currently holds data for more than 1.4 million maternal-newborn dyads, including linkable population data on the use of: all publicly funded prenatal screening modalities (including MMS and cfDNA screening); pre- and post-natal cytogenetic data from all laboratories in Ontario; and birth outcomes (born live or deceased, suspected or confirmed congenital anomalies) from every birthing hospital and midwifery practice group in the province.

### Inclusion and Exclusion Criteria

This study included all singleton pregnancies in Ontario with an EDD between September 1, 2016 and March 31, 2019 (coinciding with Ontario’s January 2016 patriation of cfDNA screening). As the objective was to measure publicly funded system performance, we excluded cfDNA screening tests that were self-paid. All eligible pregnancies from BORN Ontario were included in our analyses. In addition, we deterministically linked maternal postal code to census data to obtain maternal neighborhood income quartiles.

### Study Objectives and Outcome Measures

Our primary objective was to assess the performance of Ontario’s prenatal screening system for detection of T21 or T18. We assessed performance by sensitivity (detection rate), specificity, and screen positive rate using confirmatory diagnostic cytogenetic results. For pregnancies without a pre- or postnatal cytogenetic result, we supplemented with birth data where neither T21 nor T18 were identified in the neonatal period; this allowed the identification *all* true negatives where cytogenetic testing was not undertaken. MMS modalities, cfDNA screening conducted after a positive MMS result, and cfDNA screening done as a first-tier option were assessed separately and then combined as an overall measure of system performance. In order to form binary classification tables for performance calculations, we excluded records with non-informative results (either screening or diagnostic). This exclusion was done on a per-screening type (MMS, cfDNA test) and per-outcome (T21, T18) basis. Records were excluded from performance analysis for any of the following reasons: (1) a screening record with no result; (2) a cytogenetic result designated as mosaic, partial, uninterpretable or inconclusive; or (3) a record with no associated abnormal/normal cytogenetic outcome *and* no negative birth outcome.

Our secondary objective was to describe the impact of the contingent screening system on subsequent screening and testing for T21 by reporting the number of MMS screen-positive pregnancies that underwent either cfDNA screening or PND, as well as the number of overall screen-positive pregnancies that underwent PND. To examine the impact of publicly funded cfDNA screening on PND, we compared the number of PND for all screened pregnancies in this cohort with a historical cohort from 2012-2013.

This study was approved by the Research Ethics Boards of Children’s Hospital of Eastern Ontario (protocol 19/06PE), Ottawa Health Sciences Network (protocol 20190482-01H), and Mount Sinai Hospital (protocol 19-0181-C).

## Results

### Participants and Demographics

Of the 379,704 pregnancies that occurred between September 1, 2016 and March 31, 2019, there were 373,682 eligible singleton pregnancies. During the study period, pregnant individuals in Ontario had access to a variety of MMS modalities, including enhanced first trimester screening (49.4%), integrated prenatal screening (33.6%), first trimester screening (8.4%), maternal serum screening (6.7%), and others (2.0%). Details of test components and timelines are described in supplementary materials (Table S1 and Figure S1b).

We observed differences for pregnancies receiving first-tier cfDNA screening (compared to those having either MMS alone or MMS and cfDNA screening): maternal age was older (expected as per our contingent criteria) and income was higher. Table 1 stratifies the number of pregnancies by screening modality, and describes the maternal and fetal characteristics.

**Table 1:** Demographic data for singleton pregnancies in Ontario with an estimated due date between September 1, 2016 and March 31, 2019 (excluding those with a self-paid screening test). Eligible pregnancies with non-informative results (either screening or diagnostic) are not included. MMS: Multiple marker screening; cfDNA: cell-free DNA; EDD: estimated due date; SD: standard deviation; GA: gestational age; BMI: body mass index; Q: quintile.

|  | MMS only<br>(n= 238,538) | cfDNA only<br>(n= 5,750) | MMS + cfDNA<br>(n= 16,808) | No Screening<br>(n= 112,586) |
| --- | --- | --- | --- | --- |
| **Maternal Age at EDD**<br>**(mean, SD)** | 31.03 (4.71) | 38.85<br>(5.25) | 36.00 (5.20) | 30.35 (5.45) |
| **GA (weeks) at MMS Screening**<br>**(mean, SD)** | 12.7 (1.1) | NA | 12.70 (0.9) | NA |
| **GA (weeks) at NIPT Screening**<br>**(mean, SD)** | NA | 13.7 (1.4) | 15.6 (4.1) | NA |
| **Maternal BMI, (n, %)** |  |  |  |  |
| <18.5 | 10536 (4.4) | 175 (3.0) | 584 (3.5) | 5360 (4.8) |
| 18.5-25 | 104428 (43.8) | 2215 (38.5) | 7095 (42.2) | 47226 (41.9) |
| 25-30 | 48570 (20.4) | 1085 (18.9) | 3433 (20.4) | 23351 (20.7) |
| ≥30 | 37276 (15.6) | 908 (15.8) | 2624 (15.6) | 19353 (17.2) |
| Missing Data | 37728 (15.8) | 1367 (23.8) | 3072 (18.3) | 17296 (15.4) |
| **Smoking, n (%)** |  |  |  |  |
| Any | 12036 (5.0) | 211 (3.7) | 540 (3.2) | 11893 (10.6) |
| None | 211004 (88.5) | 4617 (80.3) | 14763 (87.8) | 92682 (82.3) |
| Missing Data | 15498 (6.5) | 922 (16.0) | 1505 (9.0) | 8011 (7.1) |
| **Income Quintile, (% column)** |  |  |  |  |
| Q1 | 51650 (21.7) | 1061 (18.5) | 3272 (19.5) | 27439 (24.4) |
| Q2 | 48620 (20.4) | 1092 (19.0) | 3183 (18.9) | 22267 (19.8) |
| Q3 | 50106 (21.0) | 1094 (19.0) | 3425 (20.4) | 21121 (18.8) |
| Q4 | 47140 (19.8) | 1123 (19.5) | 3648 (21.7) | 18626 (16.5) |
| Q5 | 36891 (15.5) | 1260 (21.9) | 3074 (18.3) | 15152 (13.5) |
| Missing Data | 4131 (1.7) | 120 (2.1) | 206 (1.2) | 7981 (7.1) |

### Screening Performance

In the 2.5 years following the patriation of two publicly funded cfDNA screening technologies to Ontario, the offer of prenatal screening was accepted for 261,096 of 373,682 singleton pregnancies (uptake of 69.9%). Of these, 255,346 (97.8%) had MMS, with 245,362 (96.1%) included in our performance analysis for T21, and 245,367 for T18. Of the 22,558 cfDNA screening records, 21,112 (93.6%) were included in our analysis for T21, and 21,115 (93.6%) for T18. For the analyses of the overall contingent program (261,096 MMS or cfDNA screening records), 250,594 (96%) were included for T21 and 250,600 (96%) for T18. Among screened pregnancies there were 2,155 tests that did not yield a result: 1,647 (0.6%) for MMS (due to presentation outside the gestational age limits) and 508 (2.3%) for cfDNA screening (test failure, including multiple attempts). Also excluded were 10,224 (3.7%) pregnancies that had prenatal screening results with an unknown outcome (i.e., they lacked both a follow-up cytogenetics result and birth outcome).

The overall sensitivity (detection rate) of Ontario’s modified-contingent prenatal screening program was 89.9% for T21 and 80.5% for T18 (Table 2). There were 4,216 screen-positive results for T21 (1.6% of all pregnancies screened) and 639 (0.2%) for T18, where cfDNA screening results are considered definitive screen results. Overall system specificity was 98.8% for T21 and 99.9% for T18, with a negative predictive value of >99.9% for both T21 and T18.

**Table 2:** Overall and modality-specific performance of a universal and publically funded prenatal screening program for trisomy 21 (T21) and 18 (T18). Through complete capture of prenatal data and birth outcomes, PSO is able to determine real-world screening performance, including true negatives (pregnancies with a negative screen result and no aneuploidy identified via PND or at birth). PSO: Prenatal Screening Ontario; MMS: multiple marker screening; cfDNA: cell-free DNA screening; PPV: positive predictive value; NPV: negative predictive value.

|  | **Overall** |  | **MMS** |  | **cfDNA** |  |
| --- | --- | --- | --- | --- | --- | --- |
|  | **T21** | **T18** | **T21** | **T18** | **T21** | **T18** |
| **Sensitivity** | 89.94<br>(87.50, 92.05) | 80.47<br>(74.53, 85.54) | 86.25<br>(83.19, 88.95) | 76.79<br>(69.66, 82.94) | 99.78<br>(98.77, 99.99) | 94.44<br>(88.30, 97.93) |
| **Specificity** | 98.76<br>(98.72, 98.81) | 99.89<br>(99.88, 99.91) | 94.99<br>(94.91, 95.08) | 99.76<br>(99.74, 99.78) | 99.81<br>(99.74, 99.87) | 99.95<br>(99.91, 99.97) |
| **PPV** | 17.26<br>(16.06, 18.51) | 39.23<br>(34.64, 43.96) | 3.94<br>(3.61, 4.29) | 17.79<br>(15.08, 20.78) | 91.99<br>(89.21, 94.24) | 90.27<br>(83.25, 95.04) |
| **NPV** | 99.97<br>(99.96, 99.98) | 99.98<br>(99.98, 99.99) | 99.97<br>(99.96, 99.97) | 99.98<br>(99.98, 99.99) | >99.99<br>(99.97, 100.00) | 99.97<br>(99.94, 99.99) |

### Impact of Contingent cfDNA Screening

We focused on T21 when examining the different prenatal screening and diagnostic testing options pursued following a screen-positive result (Figure 2). For 13,396 pregnancies with a MMS screen-positive result, 11,443 (85%) underwent follow-up testing: cfDNA screening for 9,901 (86.5%) and PND for 1,542 (13.5%); 15% had no further testing. Of the 372 pregnancies that were cfDNA screen-positive following screen-positive MMS, PND was performed for 261 (70.2%); and of the 159 pregnancies with a positive result following first-tier cfDNA screening, PND was performed for 81 (50.9%). Since the integration of publicly funded cfDNA screening in 2016, PND was performed for 6,242 pregnancies in our cohort (2.4% of all screened pregnancies; data not shown).

**Figure 2:**
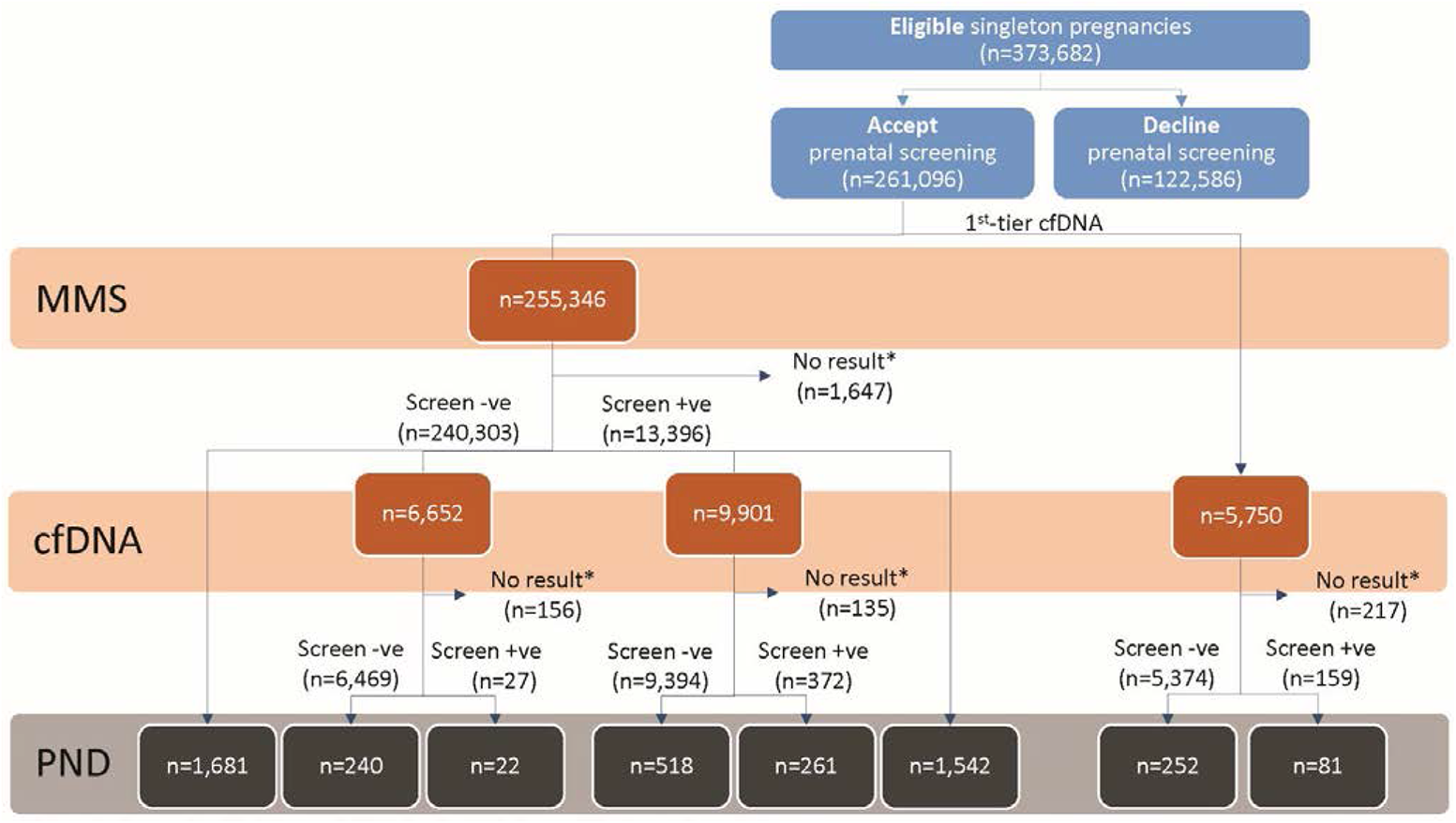
Screening Uptake and PND for Ontario’s modified-contingent cfDNA prenatal screening model for T21. During the two-year study period, an offer of prenatal screening was accepted for 261,096 singleton pregnancies (69.9%). This flowchart illustrates the variety of screening and testing options pursued by pregnant Ontarians, which models cannot capture. Real-world utilization is very different than the ideal model presented in Figure 1. MMS: maternal serum screening; cfDNA: cell-free DNA screening; PND: invasive prenatal diagnosis.

## Discussion

### Statement of Principal Findings

In our single payer health system, we were able to report on the population-based performance of contingent prenatal screening for a cohort of 373,682 pregnancies. We report an overall uptake of 69.9%; we also demonstrate a sensitivity of 89.9% for T21 and 80.5% for T18, with a screen-positive rate of 1.6% for T21 and 0.2% for T18. In agreement with other publications [27,28], our data demonstrate a first-tier cfDNA screening sensitivity of 99.8% for T21 and 94.4% for T18, with a test failure rate of 2.2% (including multiple attempts). We observed a nearly two-fold (47%) reduction in PND since the integration of cfDNA screening in 2014: 2.4% of all screened pregnancies underwent PND, compared with 4.4% (4,208 of 96,501 screened pregnancies) from a 2012-2013 cohort of 145,777 singleton pregnancies in Ontario (unpublished data). Finally, participants in Ontario’s prenatal screening program pursued a variety of options following a screen-positive result that models cannot capture. Our findings are consistent with other publications [29] that demonstrate how pregnant individuals may not pursue PND following a screen-positive result, even following a first-tier cfDNA test.

### Uptake and Performance compared with other contingent cfDNA screening systems

As with other publicly funded systems, our contingent model was chosen to provide equitable access to contingent cfDNA screening within a system that optimizes detection and reduces the overall screen-positive and PND rate, without significantly increasing overall cost [6]. Ontario’s prenatal screening system performs as expected based on our previously-modeled data [30]: we observed a slight increase in uptake (70% vs 67%) and similar performance. The contingent performance (100% sensitivity with 98.8% specificity) reported from Denmark [21] is better than that of Ontario’s overall system, largely due to Denmark’s MMS performance and screening cut-offs [19]. Each component of Ontario’s system has comparable performance with those reported from Australia (89.6% sensitivity for FTS and 100% for cfDNA screening), where cfDNA screening is offered as a self-paid test. During the study period, Ontario offered a variety of MMS modalities; a recent shift to a system predominately based on enhanced first trimester screening (eFTS) will allow for improved performance and more streamlined performance management. Improving T21 detection rates in Ontario may require lowering MMS screening cut-offs; determining the appropriate cut-offs is important in a public system, both to control costs and avoid potential harm created by false-positive results [8,17].

### Study Strengths and Weaknesses

Our study provides the largest population-based performance analysis of a contingent cfDNA prenatal screening system to date, owing to the high-quality perinatal data set available through BORN Ontario. With BORN Ontario’s complete capture of prenatal data and birth outcomes, including all pregnancies and births unaffected by aneuploidy, we were able to identify true negative cases (pregnancies without aneuploidy and with a negative screen result) and report on true overall system specificity and negative predictive value.

In our cohort, 3.7% of screened pregnancies were excluded from performance analyses because they had an unknown outcome. This proportion is lower than the general risk of miscarriage after 12w gestation (approximately 6%) [31,32]. Given that our data include cytogenetics results for PND as well as for products of conception, we speculate that most of these unknown outcomes are unrelated to aneuploidy. Nonetheless, improving the capture of such outcomes is an ongoing area of improvement.

### Implications for Clinicians and Policy Makers

Given the sample size and data quality, our study provides useful insights for other jurisdictions looking to incorporate cfDNA screening in a cost-effective manner and demonstrates the value of comprehensive, registry-based data to monitor and optimize system performance. An illustration of our ability to optimize system performance occurred at the outset of the COVID-19 pandemic: within three weeks of learning of an anticipated decrease in access to nuchal translucency scans, PSO implemented evidence-based screening algorithm adjustments. These adjustments allowed for our detection rate to be maintained even in the absence of NT, and increased access to cfDNA screening where appropriate.

Routinely-collected population data also allows for accurate tracking of test utilization as individuals navigate the program. For example, the high accuracy and non-invasiveness of cfDNA screening has led to concerns regarding the ‘routinization’ of this test and its impact on informed choice [33,34]. However, our data demonstrate that while there is considerable uptake of publicly funded cfDNA screening, only 65.2% (range 61.2-69.1%) of pregnant individuals receiving a positive result from cfDNA-screening (contingent or first-tier) proceed to PND. This supports earlier reports that pregnant individuals value the option of cfDNA screening in order to have information earlier in pregnancy, but make subsequent choices based on individual considerations [29]. It is important to note that we were unable to assess the impact of cfDNA screening on pregnancy termination rates, as BORN Ontario does not routinely collect information for these procedures.

### Future Directions

This work demonstrates the power of registry data to evaluate and optimize prenatal screening system performance. From this foundation, PSO can rapidly optimize algorithms and make evidence-based recommendations regarding the integration of new technologies (e.g., new approaches to cfDNA aneuploidy screening), targets (e.g., microdeletions), and conditions (e.g., preeclampsia, congenital anomalies). PSO is already using this real-world data to answer key questions such as the cost effectiveness of Ontario’s prenatal screening system, predictive value of no-call cfDNA screening, and the screening utility of soft markers or mosaic cytogenetic findings.

## Conclusion

We have demonstrated population-based prenatal screening performance in a publicly funded health care system, anchored by a robust comprehensive data registry. With the ability to supplement cytogenetics results with normal birth outcomes, we are able to capture all true negatives and report on complete system performance, including sensitivity, specificity, positive predictive value and negative predictive value. We analyzed 373,682 singleton pregnancies over a 2.5 year period and demonstrated that Ontario’s prenatal screening system performed as expected based on modeled data. Our contingent model was designed to provide equitable access to cfDNA screening using a system that optimizes detection, reduces screen-positives and PND, and does not significantly increase overall cost. By analyzing data in real-time, PSO can rapidly optimize performance or respond to system pressures, such as the evidence-based adjustments to screening algorithms we made in response to decreased access to nuchal translucency scans during the COVID-19 pandemic. PSO will continue to use this real-world data to improve our system to the benefit of patients, and to answer important questions about the use and utility of emerging screening methods.

## Data Availability

The data set for this study is held securely at the prescribed registry BORN Ontario. Data sharing regulations prevent these data from being made available publicly. Enquiries regarding BORN data may be directed to.

## Notes

### Competing Interest Statement

The authors have declared no competing interest.

### Funding Statement

The authors did not receive any funding for this work.

